# COVID-19 Vaccination Strategies Considering Hesitancy Using Particle-Based Epidemic Simulation

**DOI:** 10.1101/2021.09.26.21264153

**Authors:** Aknur Karabay, Askat Kuzdeuov, Huseyin Atakan Varol

## Abstract

Vaccine hesitancy is one of the critical factors in achieving herd immunity and suppressing the COVID-19 epidemic. Many countries face this as an acute public health issue that diminishes the efficacy of their vaccination campaigns. Epidemic modeling and simulation can be used to predict the effects of different vaccination strategies. In this work, we present an open-source particle-based COVID-19 simulator with a vaccination module capable of taking into account the vaccine hesitancy of the population. To demonstrate the efficacy of the simulator, we conducted extensive simulations for the province of Lecco, Italy. The results indicate that the combination of both high vaccination rate and low hesitancy leads to faster epidemic suppression.

## I. Introduction

With the rollout of a number of effective COVID-19 vaccines globally, humanity finally has the hope of suppressing the COVID-19 epidemic by achieving herd immunity through widespread vaccination. However, this prospect is threatened by vaccine hesitancy. Before the COVID-19 epidemic, prophetically, World Health Organization (WHO) pinpointed vaccine hesitancy as one of the ten global health threats to be addressed in 2019 [1]. Strategic Advisory Group of Experts (SAGE) defines vaccine hesitancy as “delay in acceptance or refusal of vaccination despite the availability of vaccination services” [2]. Vaccine hesitancy is a complex and context-dependent concept with three primary aspects: complacency, confidence, and convenience that represent an underestimation of disease-associated risks, the uncertainty of vaccine safety, and seamless access to vaccination, respectively [3]. Anti-vaccination campaigns, low level of vaccine literacy, and misinformation on social media platforms have a substantial impact on people’s vaccine perception across the globe. Association of COVID-19 with bioweapons, questioning not only the effectiveness and safety of vaccines but even whether COVID-19 exists seeded doubts on vaccination in certain socio-political groups [1], [4].

A number of surveys were performed in various countries to assess the COVID-19 vaccine acceptance. The results emphasized the importance of educating people for the success of effective vaccination campaigns. The reported acceptance rates vary across countries (94.3% in Malaysia, 91.3% in China, 53.7% in Italy, and 23.6% in Kuwait) [3]. Furthermore, lower hesitancy was observed among health-care workers and people in high-risk groups [5].

The estimates of minimum COVID-19 vaccination coverage to achieve herd immunity vary from 55% to 85% for different countries [4], [6]. Yet, the age restriction for the existing vaccines (i.e., vaccination is available only to individuals above 18 years old), as well as health restrictions on a certain portion of the eligible age groups might result in the requirement of higher vaccine coverage for the rest of the population to achieve community immunity.

Besides vaccine hesitancy, factors associated with the production, logistics, and storage of vaccines are of critical importance for predicting the epidemic dynamics and developing effective strategies to combat the pandemic. In this context, epidemic simulators are essential for finding optimal vaccination strategies considering the above-mentioned constraints and scenarios. In our previous work, we developed a vaccination simulator for COVID-19 based on a particle-based SEIR (susceptible-exposed-infected-recovered) epidemic model [7]. In this work, we extend our previous simulator for the “effective immunization” case after vaccination with the addition of the hesitancy parameter to the particle model.

## II. Methodology

In this work, we use an extension of our previous particle-based simulator with vaccination, contact tracing, and testing modules [7]. Due to the page restrictions, this section only describes the implementation of the vaccine hesitancy in the vaccination module. For further details on the simulator, the reader is referred to [7].

### A. Particle Model

In our simulator, each particle *p* represents an individual with the following attributes:

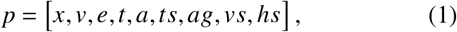

where *x* ∈ ℝ^2^ and *v* ∈ ℝ^2^ are the position and velocity of the particle; *e* is the epidemic status of the particle according to the statechart in Fig. 1; *t* is the time passed in the present epidemic state; *a* denotes whether the particle uses a contact-tracing application; *ts* denotes COVID-19 test result; *ag* indicates the age group of the particle (i.e., 1-10, 11-20 years old, etc.); *vs* and *hs* denote whether the particle is vaccinated and its vaccine hesitancy status, respectively. We use a 2D map to model the motion and the interaction of the particles. Based on the population size of the simulated region, *n* particles are randomly distributed on the map with random initial velocities with the following constraints −1 ≤ *x*_*i j*_ ≤ 1 and −*v*_*max*_ ≤ *v*_*i j*_ ≤ *v*_*max*_, where *i* = 1, …, *n* is the id of a particle, and *j* = 1, 2 denotes the dimensions on the map.

**Fig. 1:**
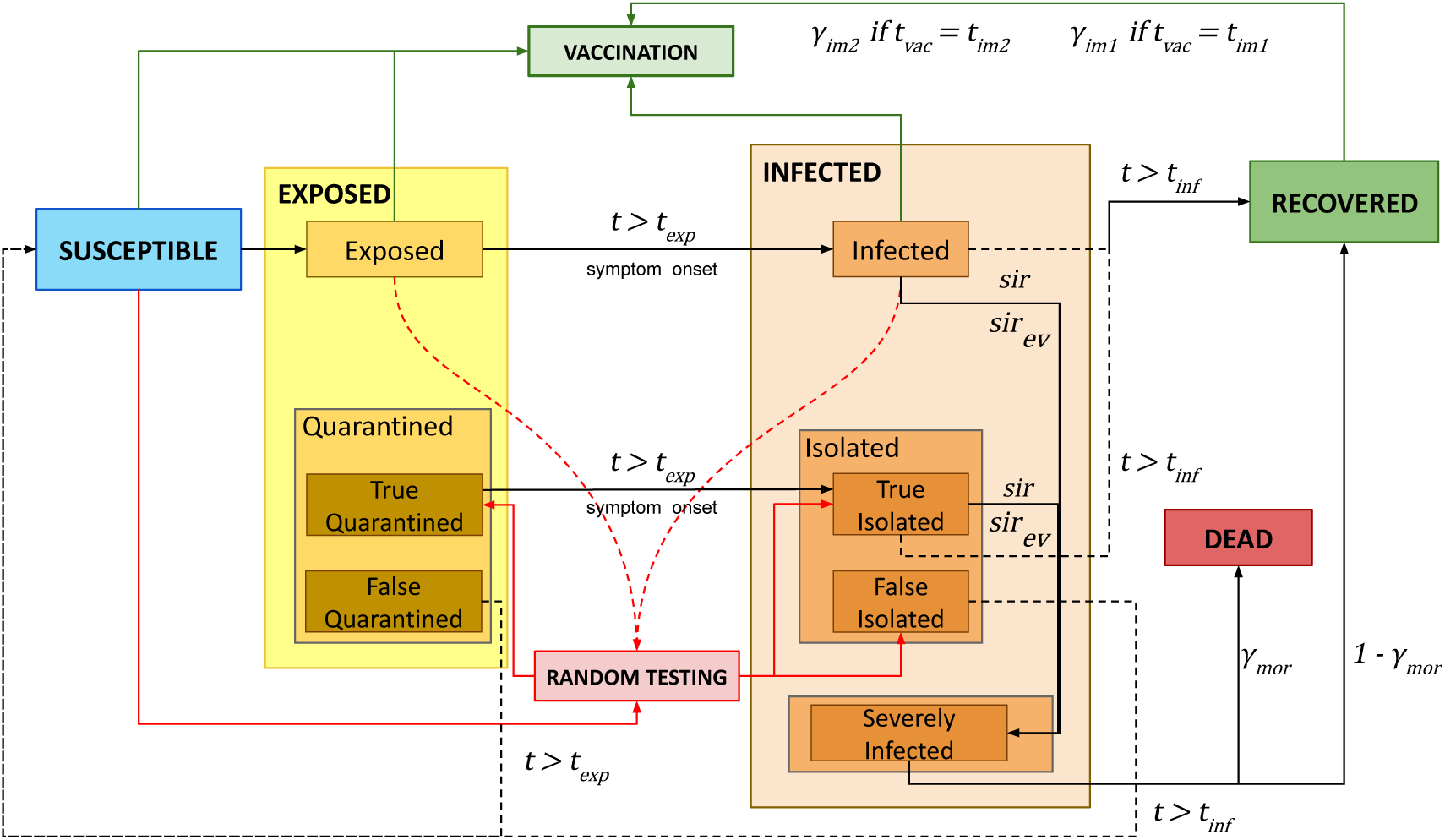
The statechart of the particle-based SEIR epidemic simulator for modeling vaccination strategies considering vaccine hesitancy rates.

At every iteration *κ* (1 ≤ *κ* ≤ *T/*Δ*t*), where *T* is the simulation length, and Δ*t* is the sampling time, the velocity *V* ∈ ℝ^*nx*2^ and position *X* ∈ ℝ^*nx*2^ of the particles are updated as:

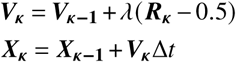

where, *λ* represents the gain that causes velocity change. *R*_***κ***_ ∈ ℝ^*nx*2^ is the vector of uniformly distributed random numbers in the range [0, 1]. This vector is normalized in the range [−0.5, 0.5] to provide a zero mean velocity change. The velocity of the particles in the quarantined, isolated, dead, and severely infected states are set to zero.

### B. Particle-based Vaccination Simulator with Hesitancy

The statechart of the particle-based SEIR simulator is illustrated in Fig. 1, and its parameters are listed in Table I. There are four main super-states: Susceptible (**S**^*s*^), Exposed (**E**^*s*^), Infected (**I**^*s*^), and Recovered (**R**^*s*^). The Exposed super-state (**E**^*s*^) has Exposed (**E**) and Quarantined (**Q**) states. The Quarantined state (**Q**) has True (**TQ**) and False (**FQ**) sub-states. The Infected (**I**^*s*^) super-state is structured in a similar way. It contains Infected (**I**), Isolated (**Iso**), and Severely Infected (**SI**) states, where the Isolated state consists of True (**TIso**) and False Isolated (**FIso**) sub-states.

**TABLE I:**
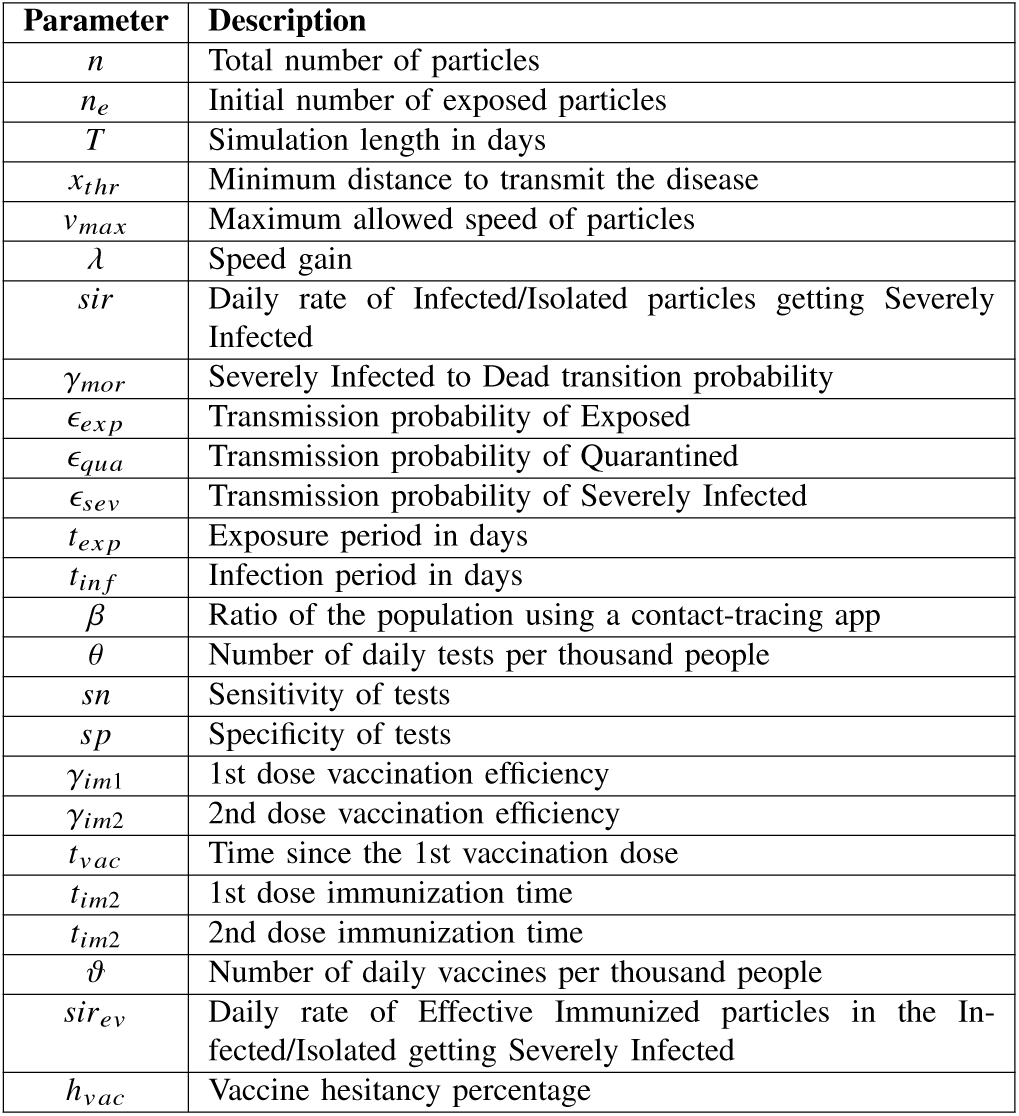
Simulation parameters and their descriptions.

Initially, all *n* particles are in the Susceptible super-state. Then, *n*_*e*_ particles are randomly selected from the **S**^*s*^ and assigned as exposed to start the epidemic. The contagious particles (from the **E, TQ, I, SI**, and **TIso** sub-states) transmit the infection within the contact threshold distance, *x*_*thr*_, and at varying disease transmission probabilities from zero to one (1 for **I**, *ϵ*_*ex p*_ for **E**; *ϵ*_*qua*_ for **TQ** and **TIso**; *ϵ*_*sev*_ for **SI**). Thus, some susceptible particles that cross the contact threshold transition to **E**. Then, after the exposure period *t*_*ex p*_, they become infected and move to state **I**. In the Infected state **I**, the particles stay for *t*_*in f*_ days. During this period, some portion of the particles transition to the Severely Infected sub-state **SI** based on the *sir* parameter, while the rest recover after *t*_*in f*_ days (and transition to the Recovered state **R**^*s*^). Meanwhile, some severely infected particles die according to the mortality rate *y*_*mor*_.

In this model, we considered “effective” post-vaccination immunization, which represents the case of a dramatic decrease in disease severity, but the reproduction and transmission of infection are enabled. Based on the vaccine hesitancy percentage, *h*_*vac*_, particles from all age groups are randomly assigned to be hesitant to the vaccine, which implies that they will not be vaccinated (*hs* is either 1 or 0 for hesitant and not hesitant, respectively). Particles from the **S**^*s*^, **R**^*s*^, **E**, and **I** states that are not marked as vaccine-hesitant go through the two-stage vaccination with a certain time period in between. The susceptible particles become immunized based on the parameters *γ*_*im*1_ at *t*_*im*1_ and *γ*_*im*2_ at *t*_*im*2_, respectively. The vaccination-immunized particles continue with the regular flow of the SEIR model once exposed to the virus, but at a much lower rate of getting severely infected, *sir*_*ev*_. While, for the particles from the **R**^*s*^ super-state, **E** and **I** states, and susceptible particles that did not gain immunity after the vaccination, the vaccine is considered to be wasted.

## III. Results

We choose the province of Lecco in Italy for a set of parametric simulations to explore the effect of vaccine hesitancy on different strategies. This province was one of the epicenters of the COVID-19 epidemic in Italy. We also used this province in our previous works [7], [8]. The simulations start from the first day of vaccination in the region (December 27, 2020). In [7], we presented the validation scenario for the province till the start of the vaccination in Italy. Therefore, we used the data from the last day of the validation scenario to initialize our simulations. The number of particles *n* was set to 337,088, i.e. the population of the province.

We ran 24 simulations for age-based and random-all strategies with the vaccine hesitancy percentage *h*_*vac*_ = {0%, 20%, 40%, 60%} and daily number of vaccines per thousand people *θ* = {2, 4, 6}. In the age-based vaccination strategy, particles are vaccinated with the oldest individuals first and then descending the age groups. While in the random-all case, particles from different age groups are vaccinated at the same time. Table II presents the number of particles to be vaccinated at each hesitancy rate, as well as the number of days to complete the vaccination considering each *θ*. The parameters used to perform the simulations are provided in Table III. As for the age-based *sir* and *sir*_*ev*_, the estimated COVID-19 age-based fatality rate for the province of Lecco was used (see Table III in [7]), and we set *sir*_*ev*_ at five percent of *sir*.

**TABLE II:** The number of vaccinated particles and vaccination period for different hesitancy rates.

| $h_{vac}$ % | Number of particles | Vaccination completed (days) | | |
| --- | --- | --- | --- | --- |
| | | $\vartheta = 2$ | $\vartheta = 4$ | $\vartheta = 6$ |
| 0 | 275,126 | 409 | 204 | 136 |
| 20 | 220,105 | 327 | 163 | 109 |
| 40 | 165,097 | 245 | 122 | 81 |
| 60 | 110,053 | 163 | 81 | 54 |

**TABLE III:** Simulation Parameters for Lecco, Italy.

| $v_{max}$ (Days) | $\lambda$ (Days) | $x_{thr}$ (Days) | $\theta$ | $sp$ | $sn$ |
| --- | --- | --- | --- | --- | --- |
| 0.007 | 0.002 | 8.6e-5 | 4 | 0.95 | 0.99 |
| $t_{inf}$ | $t_{exp}$ | $\epsilon_{exp}$ | $\epsilon_{qua}$ | $\epsilon_{sev}$ | $\gamma_{mor}$ |
| 14 | 5 | 0.7 | 0.3 | 0.3 | 0.14 |
| $\gamma_{im1}$ | $\gamma_{im2}$ | $t_{im1}$ | $t_{im2}$ | $\beta$ | $T$ (Days) |
| 52% | 95% | 12 | 28 | 0 | 420 |

The averaged results of five simulations for each scenario are presented in Figs. 2 and 3. With the increasing daily vaccination rate per thousand people, the results of the scenarios at different vaccine hesitancy rates diverge (see Fig. 2). However, the difference between the age-based and random-all strategies shrinks as both *θ* and *h*_*vac*_ grow. The comparison of different vaccination strategies at different vaccine hesitancy levels is presented in Fig. 3. At lower hesitancy, the epidemic outcome is very sensitive to the vaccination rate and selected strategy. However, as vaccine hesitancy increases, varying *θ* and strategies does not result in significant differences in the number of deaths.

**Fig. 2:**
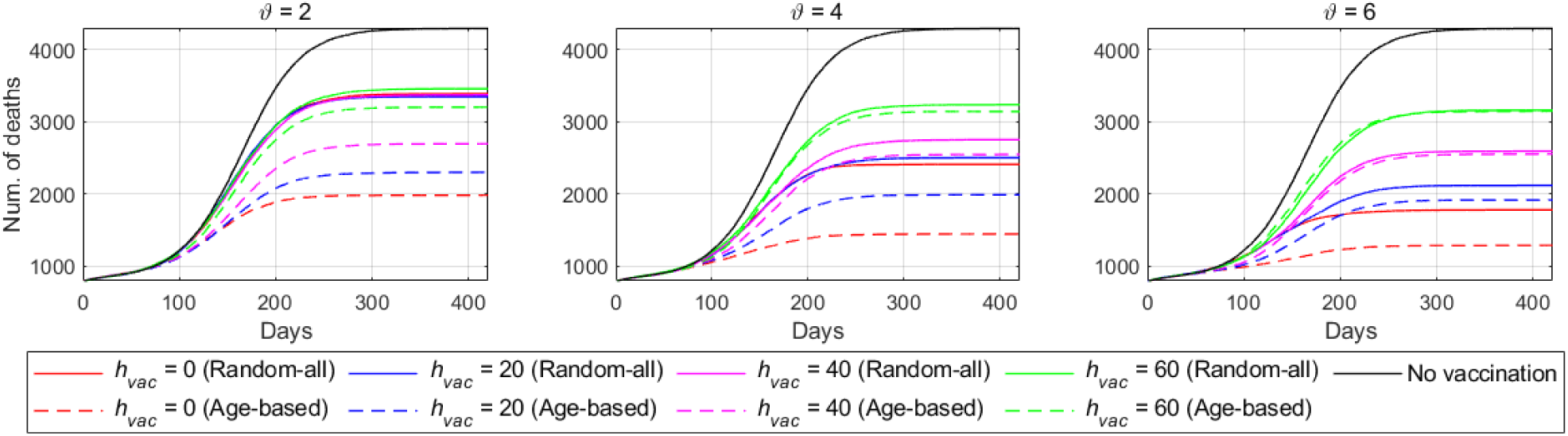
Comparison of deaths at different vaccine hesitancy levels for three daily vaccination rates per thousand people.

**Fig. 3:**
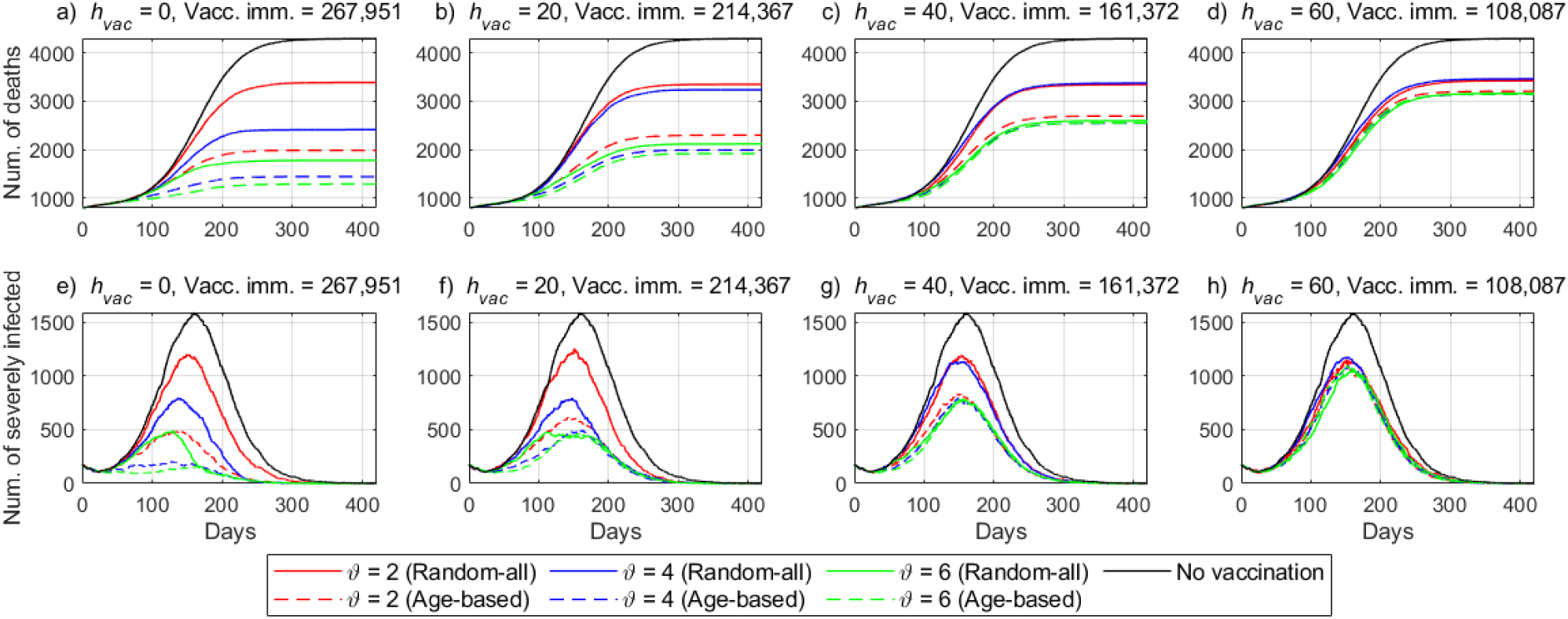
Comparison of deaths (a-d) and severely infected (e-h) for varying vaccine hesitancy levels at three daily vaccination rates.

## IV. Discussion

Overall, the age-based vaccination at highest vaccination rate and no hesitancy (*θ* = 6 and *h*_*vac*_ = 0) results in the minimum number of deaths. As shown in Fig. 3, in the case of the random-all vaccination strategy, there is a dramatic decrease in the number of deaths from *θ* = 4 to *θ* = 6 at different vaccine hesitancy rates.

In the case of the age-based vaccination strategy, the number of deaths number is reduced considerably for *θ* = 2 and *θ* = 4 for different hesitancy rates. These observations suggest that the optimal vaccination rate is higher in the random-all strategy compared to age-based.

The efficacy of the vaccination strategies at different hesitancy rates can be observed from Fig. 3. Herd immunity can be achieved rapidly with the age-based strategy with zero hesitancy and at high daily vaccination rates (*θ* = 4 and *θ* = 6). In general, a high vaccination rate with a low vaccine hesitancy has a substantial positive effect on epidemic control.

Even though the daily vaccination rate is more controllable than the vaccine hesitancy, the latter should not be under-estimated, as the epidemic dynamics and herd immunity are exceedingly sensitive to vaccine coverage. Therefore, in order to achieve an effective strategy for suppressing the epidemic, both vaccination logistics and the vaccine literacy of the society are of utmost importance.

## V. Conclusions

In this work, we presented a tool to simulate the epidemic dynamics and vaccination strategies for COVID-19 taking into account the vaccine hesitancy of the population. The simulator was written in MATLAB R2020, and the source code is available on GitHub^1^ under the MIT license.

According to the performed analysis, vaccine hesitancy is a vital factor to consider in order to deploy effective vaccination strategies and suppress the COVID-19 epidemic. The presented results have some limitations since they consider a single strain of COVID-19. As pointed out by Aschwanden [9], the delay in vaccine coverage in different countries might result in the emergence of vaccine-resistant strains of the virus, which would endanger the development of herd immunity. Noting that the vaccine supply is growing, increasing the vaccine literacy of the population might lead to a higher yield from national vaccination campaigns.

## Data Availability

N/A

https://github.com/IS2AI/Particle-Based-COVID19-Simulator/tree/Vaccine-Hesitancy-model

## Footnotes

1 https://github.com/IS2AI/Particle-Based-COVID19-Simulator/tree/Vaccine-Hesitancy-model

## References

[1] J. V. Lazarus, S. C. Ratzan, A. Palayew et al., “A global survey of potential acceptance of a COVID-19 vaccine,” Nature Medicine, vol. 27, no. 2, pp. 225–228, Feb 2021. [Online]. Available: https://doi.org/10.1038/s41591-020-1124-9

[2] N. E. MacDonald, X. Liang, M. Chaudhuri, E. Dubé, and others., “Vaccine hesitancy: Definition, scope and determinants,” Vaccine, vol. 33, no. 34, pp. 4161–4164, Aug 2015.

[3] M. Sallam, “COVID-19 vaccine hesitancy worldwide: A concise systematic review of vaccine acceptance rates,” Vaccines, vol. 9, no. 2, Feb 2021. [Online]. Available: https://pubmed.ncbi.nlm.nih.gov/33669441

[4] S. Loomba, A. de Figueiredo, S. J. Piatek, K. de Graaf, and H. J. Larson, “Measuring the impact of COVID-19 vaccine misinformation on vaccination intent in the UK and USA,” Nature Human Behaviour, vol. 5, no. 3, pp. 337–348, Mar 2021. [Online]. Available: https://doi.org/10.1038/s41562-021-01056-1

[5] A. A. Dror, N. Eisenbach, S. Taiber, N. G. Morozov, M. Mizrachi, A. Zigron et al., “Vaccine hesitancy: The next challenge in the fight against COVID-19,” European Journal of Epidemiology, vol. 35, no. 8, pp. 775–779, Aug 2020. [Online]. Available: https://doi.org/10.1007/s10654-020-00671-y

[6] K. O. Kwok, F. Lai, W. I. Wei, S. Y. S. Wong, and J. W. T. Tang, “Herd immunity - estimating the level required to halt the COVID-19 epidemics in affected countries,” Journal Infection, vol. 80, no. 6, pp. e32–e33, Jun 2020.

[7] A. Karabay, A. Kuzdeuov, M. Lewis, and H. A. Varol, “A vaccination simulator for COVID-19: Effective and sterilizing immunization cases,” medRxiv, 2021. [Online]. Available: https://www.medrxiv.org/content/early/2021/04/04/2021.03.28.21254468

[8] A. Kuzdeuov, A. Karabay, D. Baimukashev, B. Ibragimov, and H. A. Varol, “Particle-based COVID-19 simulator with contact tracing and testing,” IEEE Open Journal of Engineering in Medicine and Biology, vol. 2, pp. 111–117, 2021.

[9] C. Aschwanden, “Five reasons why COVID herd immunity is probably impossible,” Nature, vol. 591, no. 7851, p. 520—522, March 2021. [Online]. Available: https://doi.org/10.1038/d41586-021-00728-2

